# Changing epidemiology and transmission dynamics of measles in Ho Chi Minh City: a comparison of the 2018–2020 and 2024–2025 outbreaks

**DOI:** 10.64898/2026.07.27.26358995

**Authors:** Lan Truong Thi Thanh, Anh Huynh, Khang Huynh, Kieu Diem Le, Thu Pham, An Tran, Tuyen Pham Thi Kim, Thao Thanh Nguyen, Nga Le Hong, Tam Nguyen Hong, Thinh Ong, Truc Thai Thanh

## Abstract

**Introduction:** Measles remains a major public health threat worldwide. Following a resurgence of cases, Ho Chi Minh City (HCMC) experienced a severe outbreak in 2024–2025 that differed from the 2018–2020 epidemic. We compared the epidemiology and transmission dynamics of these two outbreaks to inform future surveillance and vaccination strategies.

**Methods:** Line-list data for measles cases reported in HCMC between 2018 and 2025 were extracted from the infectious disease surveillance system. Demographic changes were assessed using cross-sectional age distributions and longitudinal birth-cohort analyses. Transmission dynamics were characterized by estimating the effective reproduction number (Rt) using the Cori and Wallinga– Teunis methods. Age-stratified Who-Infected-Whom transmission matrices were constructed to quantify within- and between-age-group transmission before and after interventions.

**Results:** The 2018–2020 outbreak primarily affected children under five years of age (median age 3.75 years, IQR 0.9–9.1), whereas the 2024–2025 outbreak shifted toward older age groups (median age 8.4 years, IQR 1–14). Birth-cohort analyses identified two major immunity gaps: children born during the COVID-19 immunization disruption (2019–2023) and adolescents and young adults born before 2014 who were not eligible for the 2018 outbreak response immunization campaign. The 2024–2025 outbreak also showed broader spatial spread and a reversed geographic trajectory compared with the earlier epidemic. Transmission patterns changed substantially, with the 2018– 2020 outbreak sustained mainly by transmission among young children, whereas the 2024–2025 outbreak was driven by older age groups, with individuals >15 years acting as an important source of infection for infants.

**Conclusion:** The epidemiology of measles in HCMC has shifted from predominantly pediatric transmission to increasing involvement of adolescents and adults, reflecting accumulated immunity gaps across multiple birth cohorts. These findings suggest that age-restricted outbreak response immunization may leave residual susceptible populations that contribute to future outbreaks. Achieving sustainable measles elimination will require vaccination strategies that address historical immunity gaps across all vulnerable age groups, in addition to maintaining high routine childhood vaccination coverage.

## Introduction

Measles is a highly contagious viral disease transmitted through the respiratory route that primarily affects individuals with inadequate immunity, particularly children under five years of age.^1^ Despite the availability of safe and effective vaccines,^2^ measles remains a major public health challenge, especially in densely populated urban settings with high population mobility.^3^ Ho Chi Minh City (HCMC), Vietnam, has experienced three major measles outbreaks over the past decade (2013–2014, 2018–2020, and 2024–2025). The most recent outbreak began in May 2024, spread rapidly, and placed considerable pressure on the healthcare system.

The 2018–2020 outbreak predominantly affected unvaccinated or partially vaccinated children. Although first-dose measles vaccine coverage reached 96%, second-dose coverage remained at only 80%, below the 95% threshold required for elimination, creating substantial immunity gaps.^4^ An outbreak response immunization (ORI) campaign targeting children aged 1–5 years subsequently achieved approximately 85% coverage.^4^ In contrast, the 2024–2025 outbreak occurred after COVID-19-related disruptions to routine immunization, which reduced community immunity among young children. The subsequent ORI campaign expanded eligibility to children aged 6– 10 years and achieved near-universal coverage.^5^ These differences in population immunity and vaccination strategies may have altered transmission patterns between the two outbreaks.

The 2024–2025 outbreak was the largest recorded in HCMC, with more than 9,000 reported cases and seven deaths.^6^ Compared with previous outbreaks, it showed marked differences in age distribution, geographic spread, and the timing of public health interventions. However, evidence describing measles transmission dynamics in Vietnam remains limited. We therefore compared the epidemiology and transmission dynamics of the 2018–2020 and 2024–2025 outbreaks in HCMC using effective reproduction number (Rt) estimates and age-specific transmission analyses to provide evidence for improving surveillance and guiding future outbreak response.

## Method

### Study design

This retrospective observational study was conducted based on the records of measles cases reported to the Ho Chi Minh City Center for Disease Control (HCDC), Vietnam, between 2018 and 2025. The linelist include all of measles cases confirmed by laboratory or clinical in Ho Chi Minh City. The diagnosis of the case followed the guidelines for the diagnosis and treatment of measles issued by the Ministry of Health, Vietnam^7^:

#### Suspected case

A patient presenting with fever and a maculopapular rash, accompanied by at least one of the following clinical signs: cough, coryza, conjunctivitis, lymphadenopathy (cervical, occipital, or post-auricular), or arthralgia/joint swelling.

#### Laboratory-confirmed case

A suspected case with a definitive diagnosis of measles confirmed by at least one of the following laboratory assays: detection of measles-specific IgM antibodies via enzyme-linked immunosorbent assay (ELISA), identification of measles-specific viral RNA segments via polymerase chain reaction (PCR), or successful viral isolation.

#### Epidemiologically linked case

A suspected case lacking clinical specimen collection but possessing a clear epidemiological link to a laboratory-confirmed or previously epidemiologically linked measles case. An epidemiological link is established by direct or potential contact within the same spatiotemporal setting, provided that the interval between the dates of rash onset for the two cases is between 7 and 21 days.

#### Discarded case

A suspected case with an adequate clinical specimen that tests negative for both measles, or a case with a definitive diagnosis of another disease.

### Inclusion and exclusion criteria

The study included measles cases recorded in the surveillance system at the Ho Chi Minh City Center for Disease Control (HCDC) from January 1st, 2018, to December 3rd, 2025. Inclusion criteria required cases to have a residential address within Ho Chi Minh City and a documented date of admission. Cases with a final classification of “discarded measles” were excluded from the analysis.

### Data collection

Data for this study were extracted from the national infectious disease surveillance system managed by the Ministry of Health (MoH), Vietnam. Case reporting from healthcare facilities to this centralized platform is conducted via two primary mechanisms: facilities equipped with proprietary electronic medical record (EMR) systems transmit data automatically through integrated interoperability platforms, whereas facilities without such systems perform manual data entry directly into the MoH web-based portal. Consequently, all notified cases across different clinical settings are consolidated into a unified national database, from which the study’s linelist was exported. The extracted data collection encompasses all cases with recorded hospital admission dates between January 1st, 2018, and December 3rd, 2025. The dataset includes the following standardized demographic and clinical variables, routinely collected by medical staff during patient intake:

#### Date of birth

Extracted from the patient’s administrative records to calculate the precise age at the time of infection, enabling age-stratified and cohort-tracking analyses.

#### Gender

The biological sex of the patient (male or female).

#### District

The patient’s current residential address at the district level within Ho Chi Minh City, utilized for spatial distribution mapping and calculating regional cumulative incidence.

#### Admission date

The exact date the patient presented and was registered at the healthcare facility. This variable serves as the primary temporal metric for constructing epidemic curves and calculating the transmission matrix and effective reproduction number (R_t_).

#### Status (inpatient/outpatient)

The clinical management setting assigned upon intake, reflecting the initial assessment of disease severity.

#### Vaccination status

The patient’s history of measles immunization (e.g., unvaccinated, number of doses received), ascertained through official immunization records (vaccination cards or national digital logs) or, when records are unavailable, through parental/self-recall.

#### Diagnostic classification

The case definition status assigned by clinicians or preventive medicine centers. This variable is crucial for data processing, as it categorizes cases into laboratory-confirmed, epidemiologically linked, suspected, or discarded cases. In this study, this field was utilized as the primary filter to identify eligible cases and systematically exclude cases with a final classification of “discarded measles.”

### Statistical analysis

Descriptive statistics were utilized to summarize the temporal and demographic characteristics of the measles outbreaks. Epidemic curves were constructed based on the dates of hospital admission. To characterize demographic shifts, age distributions were analyzed using both traditional cross-sectional proportions and a birth-cohort tracking approach, enabling the longitudinal observation of specific generational susceptibilities across the two distinct outbreaks. Spatial dissemination was evaluated by calculating the cumulative incidence per 100,000 population at the district level, mapped across six equal temporal phases for each outbreak to visualize geographic expansion.

Transmission dynamics were quantified by estimating the effective reproduction number (R_t_) using two distinct methodological frameworks. First, the backward-looking method proposed by Cori et al. was applied using a 14-day sliding window to provide real-time R_t_ estimates based on the incidence time series. Second, the forward-looking case reproduction number (R_t_) framework by Wallinga and Teunis was implemented to reconstruct individual transmission trees. This method utilizes the probability distribution of the serial interval, which was modeled following a Gamma distribution with a mean of 14.5 days and a standard deviation of 3.25 days. The Wallinga and Teunis approach permitted high-resolution stratification of R_t_ by specific age groups and districts over time. To statistically evaluate the differences in the daily R_t_ estimates derived from these two methodologies, a paired Wilcoxon signed-rank test was employed.

Furthermore, the probabilistic transmission network derived from the Wallinga and Teunis method was aggregated to construct 5×5 “Who-Infected-Whom” transmission matrices. These matrices quantified the inter- and intra-cohort infectious pressure (F0 to F1) among five age strata (<1, 1–5, 6–10, 11–15, and >15 years) across distinct pre- and post-intervention phases.

All statistical analyses, mathematical modeling, and spatial data visualizations were performed using R software (R Foundation for Statistical Computing, Vienna, Austria). The *EpiEstim* package was utilized for the Cori method. R_t_ estimations, while spatial analyses and data manipulation were conducted using the *sf* and *tidyverse* suites, respectively. Graphical outputs were generated utilizing *ggplot2* and *patchwork*. To ensure full methodological transparency and computational reproducibility, the complete analytical code and anonymized datasets have been made publicly available in a GitHub repository at https://github.com/AnhsHuynh/measles-outbreak.

### Estimation of the Effective Reproduction Number (R_**t**_**) by Cori et al**^8^

Data Pre-processing: The daily incidence time series (I_t_) was cleaned and zero-filled for days with no reported cases to ensure temporal continuity. Serial Interval (SI) distribution: We assumed a Gamma distribution of serial interval based on established international literature for measles, utilizing a mean SI of 14.5 days and a standard deviation of 3.25 days.^10^

(1)

(2) Bayesian Estimation Framework: R_t_ was estimated over a 14-day sliding window to stabilize the daily fluctuations. The posterior distribution of R_t_ at time (*t*) was derived assuming a Gamma prior distribution (parameters *a, b*) updated by the likelihood of the observed incidence data (I_t_) and the renewal equation (Λ_t_).

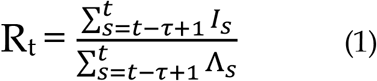

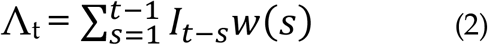

To capture spatial heterogeneity in transmission dynamics, R_t_ was estimated independently for each of the 22 districts. Given that sparse daily case counts at the district level can introduce significant stochastic noise into instantaneous estimates, incidence data were aggregated by epidemiological week. Zero-filling method was subsequently applied to non-reporting weeks to maintain the temporal continuity necessary for stable model estimation.

### Estimation of the Effective Reproduction Number (R_**t**_**) by Wallinga and Teunis**^10^

The methodology proposed by Wallinga and Teunis (2004) provides a cohort-based, forward-looking approach to estimating the effective reproduction number. In contrast to retrospective (backward-looking) methods like Cori et al., which rely on the ratio of current to past incidence, the Wallinga and Teunis framework reconstructs transmission networks. It estimates the mathematical probability that a given primary case *j* infected a secondary case *i* based on the probability density function of the generation time or serial interval.

The probability p_ij_ that case *i* was infected by case *j* is calculated as:

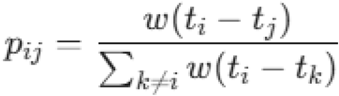

Where:

- t_i_, t_j_: The dates of symptom onset (or hospital admission) for cases $i$ and $j$, respectively.
- w(Δt): The probability density function of the serial interval (SI), which is typically assumed to follow a Gamma distribution.
- The denominator represents the sum of the probabilities that case *i* was infected by any other preceding case *k* within the network.

Subsequently, the individual reproduction number for case j (R_j_) is defined as the sum of all infection probabilities attributed to that specific individual:

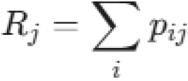

The effective reproduction number at time *t* (R_t_) is then computed by averaging the R_j_ values of all incident cases that occurred on day *t*:

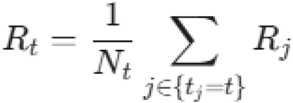

## Results

The epidemic curves (Figure 1) summarize the overall progression of the two outbreaks. The 2024–2025 outbreak had a longer growth phase before reaching its peak (7.3 months) than the 2018–2020 outbreak (5.1 months), although the overall outbreak durations were similar (18.2 vs. 19.1 months). The overall effective reproduction number (R_t_) (Figure 5) further illustrates these differences in transmission dynamics. Both outbreaks began with high R_t_ values that declined following implementation of outbreak response immunization. In 2018– 2020, R_t_ decreased rapidly and remained close to or below one for most of the remaining outbreak. In contrast, the 2024–2025 outbreak exhibited greater temporal variability, with repeated increases in R_t_ and several secondary peaks despite the vaccination campaign, indicating more sustained and heterogeneous transmission before the outbreak was controlled.

**Figure 1.**
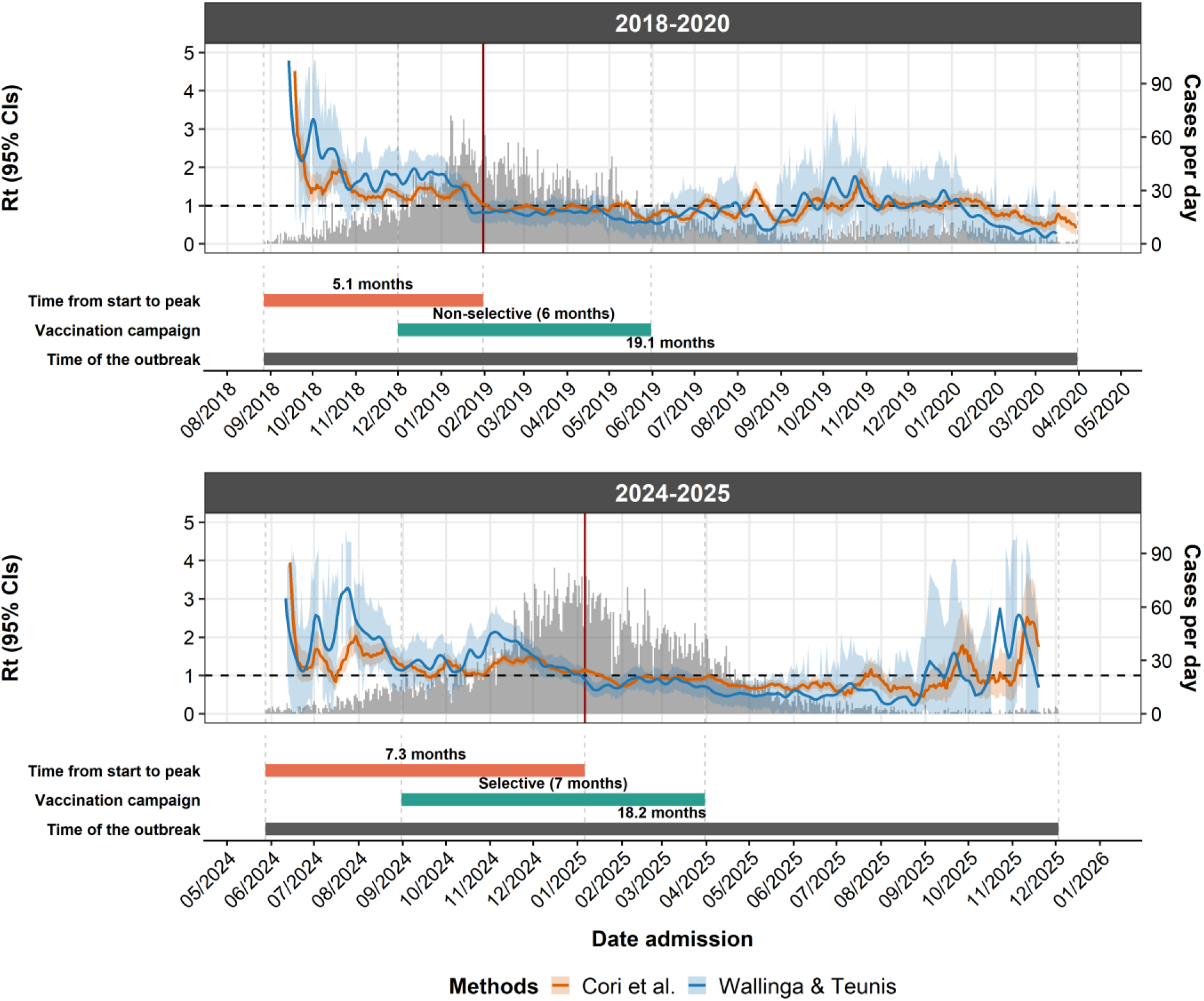
Temporal dynamics of daily incidence and Rt for measles between the 2018-2020 and 2024-2025 outbreak in Ho Chi Minh City. Daily reported cases are shown as grey bars (right y-axis). Rt was estimated using the Cori et al. (orange) and Wallinga & Teunis (blue) methods; shaded areas represent 95% confidence intervals. The dashed horizontal line indicates Rt = 1, and the vertical red line marks the epidemic peak. The lower panel indicates the time from outbreak onset to the epidemic peak, the duration and type of outbreak response immunization campaign, and the total outbreak duration.

**Figure 2.**
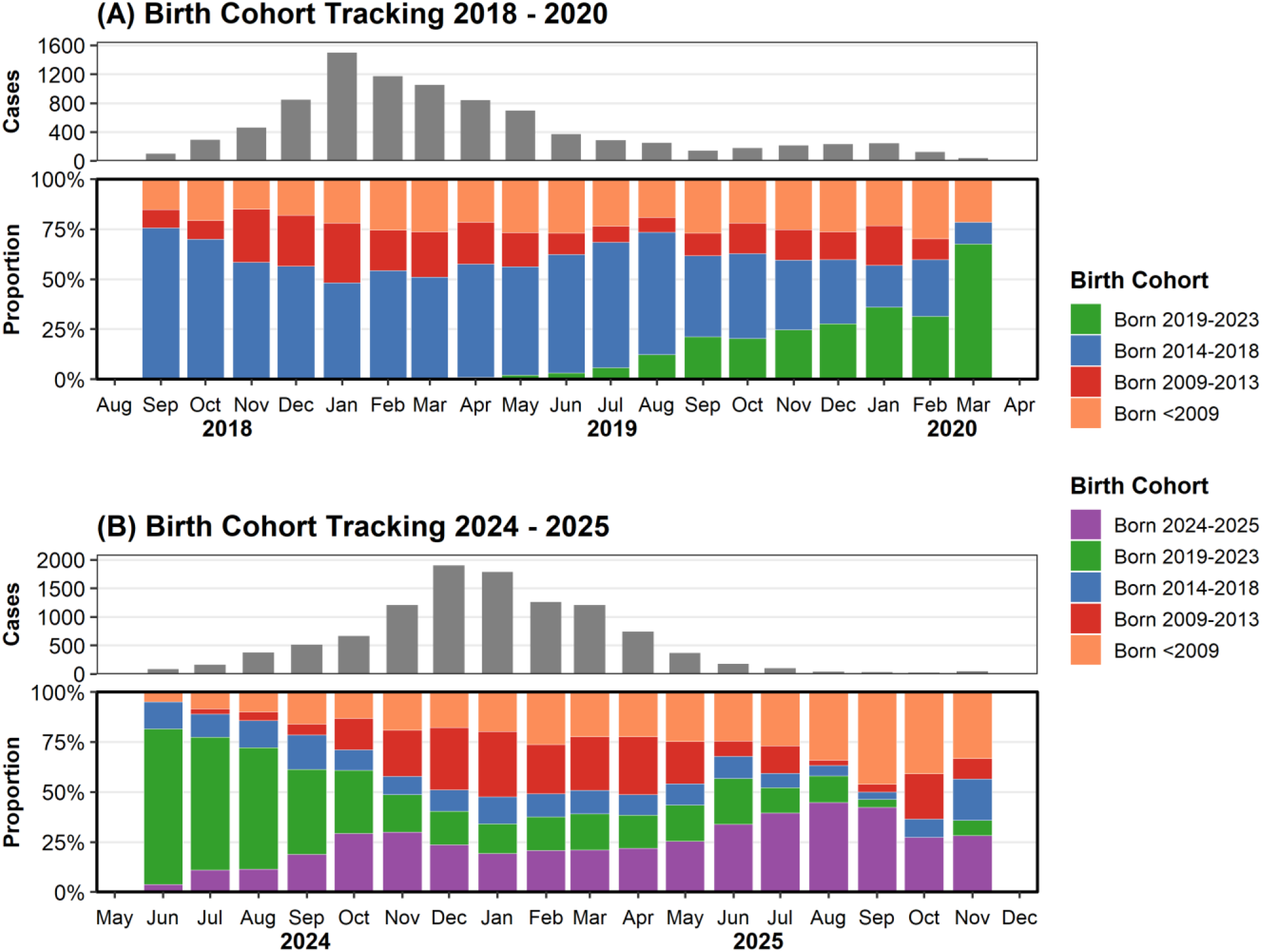
Birth cohort tracking of measles cases during the 2018–2020 and 2024–2025 outbreaks in Ho Chi Minh City. Grey bars show the monthly number of cases, and stacked bars show the monthly proportion of cases by birth cohort

Table 2 compares the demographic and clinical characteristics of measles cases in the 2018– 2020 and 2024–2025 outbreaks. The median age increased from 3.75 years (IQR 0.9–9.1) in 2018–2020 to 8.4 years (IQR 1–14) in 2024–2025 (p<0.001), reflecting a clear shift toward older age groups. In the 2018–2020 outbreak, most cases occurred among children aged 1–5 years and 6–10 years, whereas in 2024–2025 a much larger proportion occurred among adolescents aged 11–15 years. The sex distribution was similar in both outbreaks, with a slight male predominance in each period (p=0.033). Most cases were hospitalized in both outbreaks, and the proportions were slightly higher in 2024–2025. Vaccination history also differed markedly: a substantially larger proportion of cases in 2024–2025 had documented prior vaccination, while the proportion with unknown status was lower than in 2018–2020 (p<0.001).

**Table 2.** Epidemiological characteristics of measles cases in Ho Chi Minh City during the outbreaks 2018-2020 and 2024-2025.

|  | 2018 – 2020<br>(n= 8,914) | 2024 – 2025<br>(n= 9,740) | p |
| --- | --- | --- | --- |
| **Age** |  |  |  |
| Year | 3.75 (0.9–9.1) | 8.4 (1–14) | <0.001<sup>(a)</sup> |
| **Age group** |  |  |  |
| <1 years | 2,339 (26.2%) | 2,363 (24.3%) |  |
| 1–5 years | 2,701 (30.3%) | 1,646 (16.9%) |  |
| 6–10 years | 1,828 (20.5%) | 1,237 (12.7%) | <0.001<sup>(b)</sup> |
| 11–15 years | 283 (3.2%) | 2,413 (24.8%) |  |
| >15 years | 1,763 (19.8%) | 2,081 (21.4%) |  |
| **Gender** |  |  |  |
| Male | 4,874 (54.7%) | 5,477 (56.2%) |  |
| Female | 4,040 (45.3%) | 4,263 (43.8%) | 0.033<sup>(b)</sup> |
| **Status** |  |  |  |
| Inpatient | 4,876 (54.7%) | 5,545 (56.9%) |  |
| Outpatient | 4,037 (45.29%) | 4,186 (43%) | <0.001<sup>(b)</sup> |
| Mortality | 1 (0.01%) | 7 (0.08%) |  |
| Others | 0 (0%) | 2 (0.02%) |  |
| **Vaccination** |  |  |  |
| Yes | 69 (0.8%) | 3,000 (30.8%) |  |
| No | 5,084 (57%) | 3,985 (40.9%) | <0.001<sup>(b)</sup> |
| Unknown | 3,761 (42.2%) | 2,755 (28.3%) |  |

Continuous variable show as median (Q1–Q3); Categorical variable show as N(%)
<sup>(a)</sup> Using the Wilcoxon’s test; <sup>(b)</sup> Using Chi-squared test

In 2018–2020, cases increasingly concentrated in children born after the June 2019 campaign, indicating that later-born cohorts became more prominent as the outbreak progressed. In 2024–2025, the outbreak was initially driven by the 2019–2023 birth cohort, which accounted for about 70–80% of cases in June–August 2024. Its contribution declined after the selective vaccination campaign began in September 2024, although transmission persisted across multiple older cohorts into late 2025.

Spatial mapping of cumulative incidence (Figure 3) showed different geographic spread patterns in the two epidemics. The 2018–2020 outbreak began in the western region and expanded eastward, with the western and southern peripheral districts most affected at the peak and the western districts remaining the hardest hit by the end of the outbreak (approximately 161 per 100,000 population). In contrast, the 2024–2025 outbreak originated mainly in the eastern region and spread westward, with the eastern districts bearing the highest burden at the peak and by the end of the outbreak (approximately 154 per 100,000 population).

**Figure 3.**
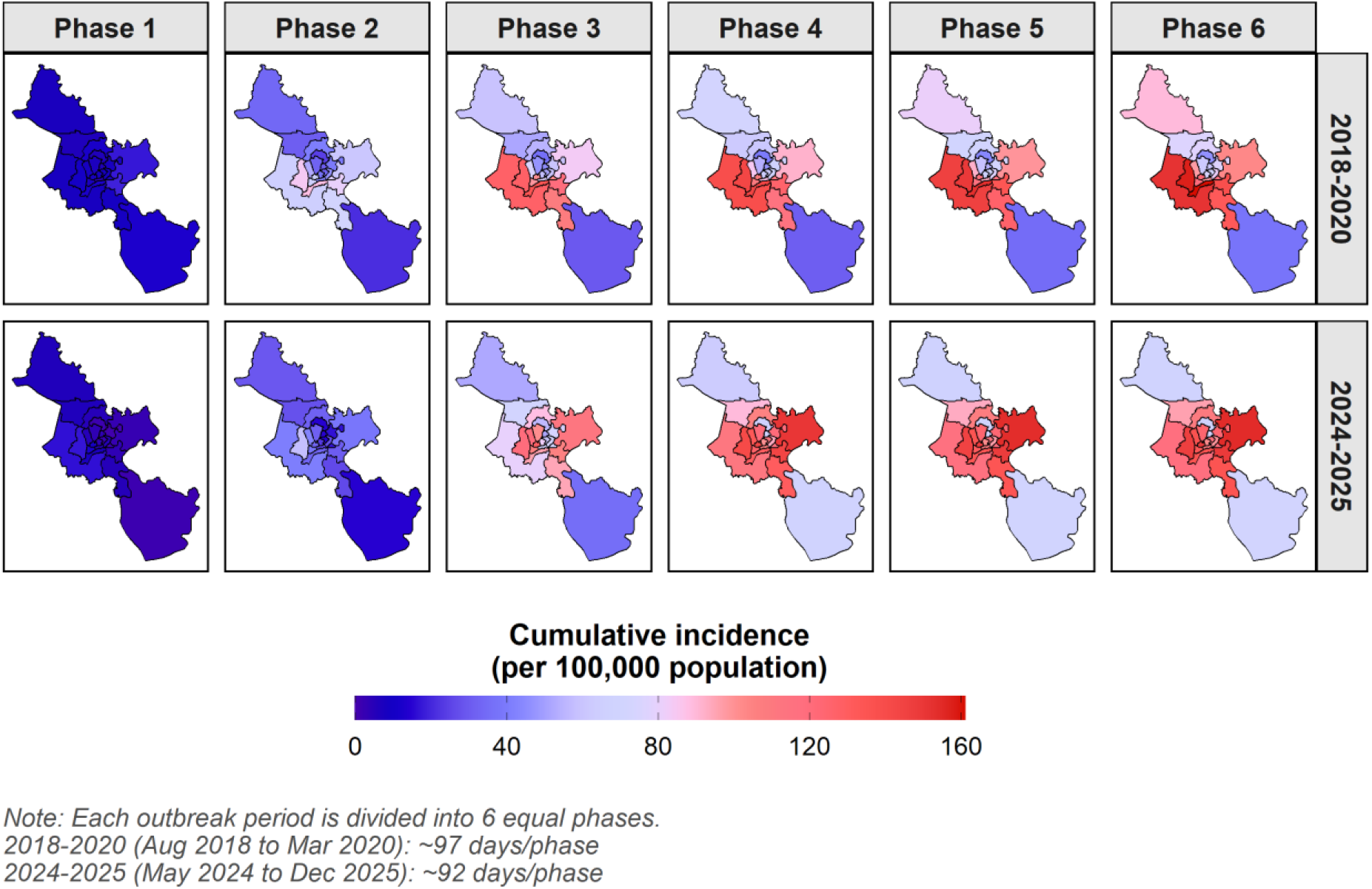
Cummulative incidence per 100.000 populations by districts

The transmission matrices (Figure 4) showed a marked shift in infection patterns between the two outbreaks. During the 2018–2020 outbreak, the highest pre-intervention transmission occurred from children aged 6–10 years to those aged 1–5 years, with Rt reaching 0.46. After the vaccination campaign, this transmission declined to 0.29. In contrast, the 2024–2025 outbreak was driven mainly by older age groups, with individuals aged >15 years acting as the main source of infection. The highest pre-intervention transmission was from those aged >15 years to infants aged <1 year, with Rt reaching 0.61, and this decreased to 0.24 after the vaccination campaign.

**Figure 4.**
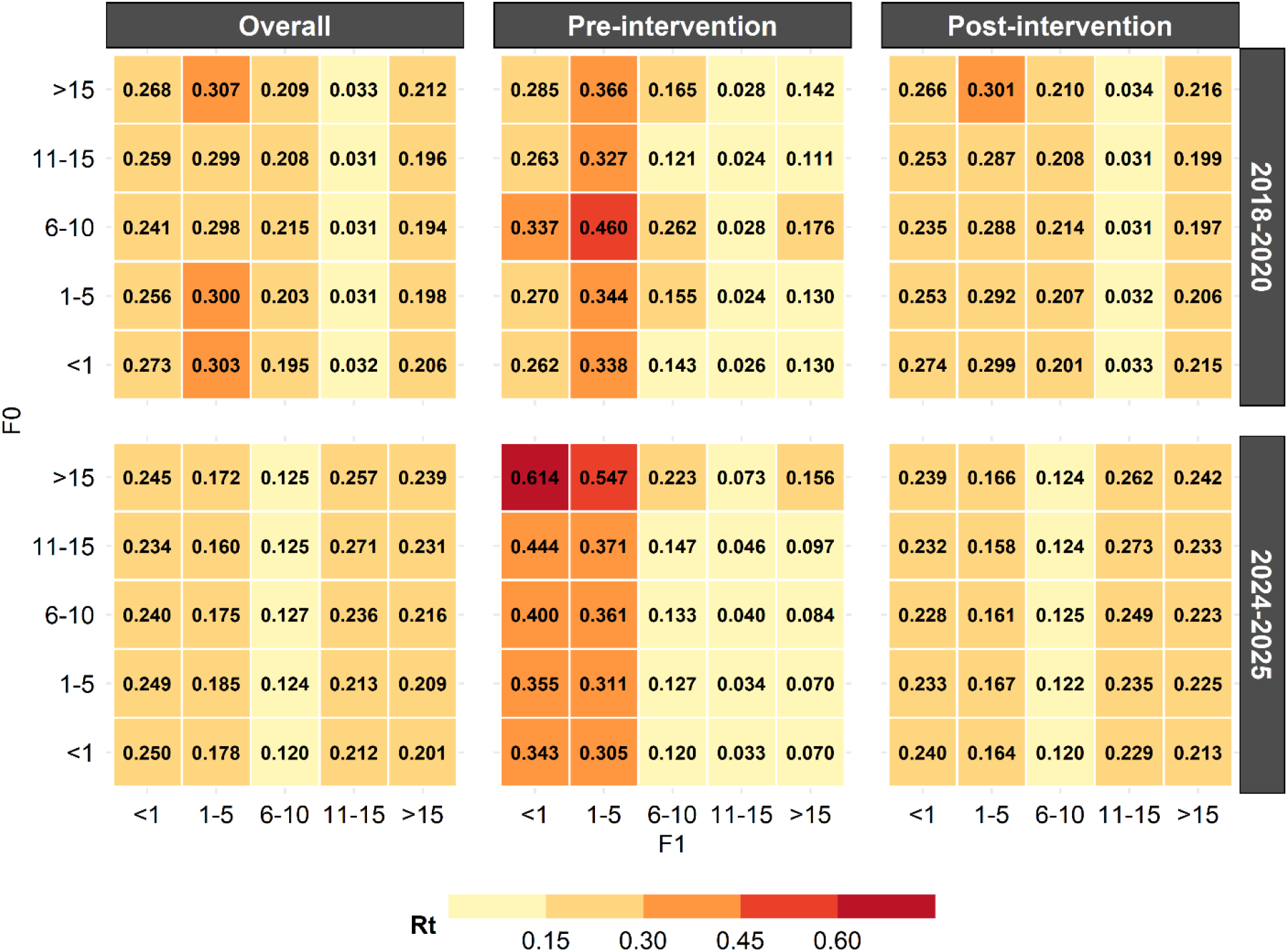
Age-stratified who-infected-whom transmission matrices for the 2018–2020 and 2024–2025 measles outbreaks in Ho Chi Minh City. Heat maps show the Rt for transmission from infector age group (F0, rows) to recipient age group (F1, columns) for the overall outbreak period, and separately for the pre-intervention and post-intervention phases. Darker colors indicate higher Rt values.

## Discussion

This study compared the epidemiology and transmission dynamics of two major measles outbreaks in Ho Chi Minh City. We observed substantial changes in the age distribution, geographic spread, and transmission patterns between the 2018–2020 and 2024–2025 outbreaks. Whereas the earlier outbreak was characterized by transmission predominantly among young children, the recent outbreak increasingly involved adolescents and adults. These findings highlight how historical vaccination strategies and COVID-19-related disruptions to routine immunization can reshape population immunity and alter the epidemiology of future outbreaks.

The age distribution of cases shifted from predominantly young children in 2018–2020 to adolescents and adults in 2024–2025, consistent with trends reported in countries with historically high vaccination coverage.^11^. Although routine measles vaccination coverage in Vietnam has remained above 90% since 2014, apart from an 81–89% decline during the COVID-19 period (2020–2023),^12^ the accumulation of susceptible adolescents and adults may reflect waning vaccine-induced immunity.^13^ This finding has important implications for measles elimination, as closing immunity gaps in older age groups may require booster vaccination. Previous modelling studies suggest that a third measles-containing vaccine dose at 18–20 years of age could help reduce these gaps and support elimination efforts.^14^

A major finding of this study is the long-term impact of ORI on future susceptibility. Birth-cohort tracking showed that children targeted by the 2018 ORI campaign remained relatively protected during the 2024–2025 outbreak, whereas those who were not eligible for vaccination aged into the adolescent and young adult groups that contributed substantially to transmission. These findings suggest that although age-targeted ORI effectively protects vaccinated cohorts, it may also leave residual susceptible populations that become apparent years later as unvaccinated cohorts age. Future outbreak responses should therefore consider historical immunity profiles in addition to the age groups most affected during the current epidemic.

The 2024–2025 outbreak also highlighted the consequences of COVID-19-related disruptions to routine immunization. The 2019–2023 birth cohort accounted for most cases at the beginning of the outbreak before declining rapidly following the vaccination campaign, demonstrating the effectiveness of catch-up vaccination in this group. However, sporadic cases also occurred among cohorts that had previously been protected by ORI. Ho Chi Minh City experiences substantial inward migration, and children moving into the city may not have benefited from previous local vaccination campaigns. This continual introduction of susceptible individuals may contribute to persistent transmission despite high reported vaccination coverage and emphasizes the importance of maintaining catch-up vaccination among mobile populations.

Changes in population immunity were reflected in the transmission patterns. During the 2018–2020 outbreak, the strongest transmission occurred between school-aged children and preschool children, consistent with the typical epidemiology of measles in populations with immunity gaps concentrated among young children. In contrast, the 2024–2025 outbreak was characterized by transmission from adults to infants, indicating that adolescents and adults had become an important reservoir of infection. This demographic shift is likely to have contributed to the broader geographic spread observed during the recent outbreak, as older individuals generally have greater mobility and more diverse social contacts than young children.

The different transmission patterns were accompanied by distinct epidemic trajectories. Although both outbreaks lasted for a similar duration, the 2024–2025 outbreak exhibited a longer growth phase and more variable effective reproduction numbers before transmission was controlled. These findings suggest that transmission became more heterogeneous, with sustained spread across multiple age groups and geographic areas. The rapid decline in cases among children following the expanded vaccination campaign demonstrates the effectiveness of selective ORI in reducing susceptibility in targeted groups. Nevertheless, continued transmission among older age groups indicates that interrupting transmission may become increasingly difficult when substantial immunity gaps accumulate outside the traditional pediatric population.

The consistent findings obtained using both the Cori and Wallinga–Teunis methods strengthen confidence in the observed transmission dynamics. The two approaches provide complementary perspectives on epidemic spread. The forward-looking Wallinga–Teunis method is sensitive to emerging transmission and useful for reconstructing transmission patterns retrospectively, whereas the backward-looking Cori method provides a more stable estimate of real-time transmissibility and is better suited for monitoring intervention effects during an outbreak. The concordance between these methods supports the robustness of our findings.

This study has some limitations. First, surveillance data likely underestimate the true burden of infection because mild or subclinical infections, particularly among partially immune adults, may not present for medical care. Second, vaccination history was incomplete for some cases and relied partly on parental or self-report when electronic records were unavailable, introducing the potential for misclassification. Third, effective reproduction numbers were estimated using hospital admission dates rather than symptom onset because of incomplete onset data. Although this approach may shift Rt estimates temporally, it is unlikely to affect comparisons between outbreaks. Finally, the Who-Infected-Whom matrices are based on probabilistic transmission inferred from serial intervals rather than confirmed transmission chains. Nevertheless, these methods remain appropriate for describing transmission patterns at the population level.

Our findings have important implications for measles surveillance and outbreak response. Maintaining high routine childhood vaccination coverage remains essential but is unlikely to be sufficient in settings where historical immunity gaps have accumulated. Periodic serological surveys could help identify susceptible cohorts before outbreaks occur, while catch-up vaccination strategies should consider both historical vaccination campaigns and changing demographic patterns. As measles epidemiology continues to evolve, outbreak response immunization may need to extend beyond young children to include susceptible adolescents and adults in order to interrupt transmission and protect infants who are too young to be vaccinated.

## Conclusion

The epidemiology of measles in Ho Chi Minh City has changed substantially, with transmission shifting from predominantly young children to adolescents and adults. This transition reflects the combined effects of historical outbreak response immunization and COVID-19-related disruptions to routine vaccination, which together reshaped population immunity across successive birth cohorts. While outbreak response immunization remains an effective tool for rapidly reducing susceptibility in targeted groups, future strategies should account for historical immunity gaps across all vulnerable age groups rather than focusing solely on children. Integrating routine surveillance with periodic assessment of population immunity and age-appropriate catch-up vaccination may improve the long-term resilience of measles control programmes and support progress toward sustainable measles elimination.

## Data Availability

The data used in this study are available from the authors and Ho Chi Minh City Center for Disease Control upon reasonable request, subject to data confidentiality restrictions

## Acknowledgements

We sincerely thank the Department of Infectious Disease Control and Prevention at the Ho Chi Minh City Center for Disease Control (HCDC) for providing the surveillance data used in this study.

## Contribution

AH, TO and LTTT developed the study concept and design. AH, LTTT, NLH and TNH coordinated the overall research process. AT, TP, KDL, KH, TNT were responsible for data extraction and conducted the scoping review. AH, AT, KDL, KH, TP conducted statistical analysis and interpreted the data. All authors contributed to drafting the manuscript, critically revised its content, and approved the final version for submission. This study was conducted under the supervision of TTT, TO.

## Ethics declarations

The research has been approved in terms of medical ethics in research from the Ethics Council in Biomedical Research of the Ho Chi Minh City Center for Disease Control (IRB number: 02/GCN-HĐĐĐ-TTKSBT, sign: November 17, 2025).

## References

1. Hübschen JM, Gouandjika-Vasilache I, Dina J. Measles. The Lancet. 2022;399(10325):678–690. doi:10.1016/S0140-6736(21)02004-3

2. Griffin DE, Lin WH, Pan CH. Measles virus, immune control, and persistence. FEMS Microbiol Rev. 2012;36(3):649–662. doi:10.1111/j.1574-6976.2012.00330.x

3. Bischops AC, Hulland EN, Patzakis M, Majumder MS. Will the USA lose its measles elimination status? The Lancet. 2026;407(10540):1679–1681. doi:10.1016/S0140-6736(26)00466-6

4. Cục Phòng bệnh. Vận động phụ huynh tiêm chủng vắc - xin phòng bệnh sởi cho trẻ đầy đủ - Trang chủ - Cổng thông tin Bộ Y tế. Accessed September 4, 2025. https://vncdc.gov.vn/van-dong-phu-huynh-tiem-chung-vac-xin-phong-benh-soi-cho-tre-day-du-nd14901.html

5. Ong T, Thuy CT, Tam NH, et al. Detection of Immunity Gap before Measles Outbreak, Ho Chi Minh City, Vietnam, 2024 - Volume 31, Number 10—October 2025 - Emerging Infectious Diseases journal - CDC. Published online October 2025. doi:10.3201/eid3110.250234

6. Gostic KM, McGough L, Baskerville EB, et al. Practical considerations for measuring the effective reproductive number, Rt. PLOS Comput Biol. 2020;16(12):e1008409. doi:10.1371/journal.pcbi.1008409

7. Bộ Y tế. Quyết định 4845/QĐ-BYT về “Hướng dẫn giám sát phòng chống bệnh Sởi Rubella.” 2012.

8. Cori A, Ferguson NM, Fraser C, Cauchemez S. A New Framework and Software to Estimate Time-Varying Reproduction Numbers During Epidemics. Am J Epidemiol. 2013;178(9):1505–1512. doi:10.1093/aje/kwt133

9. Worden L, Ackley SF, Zipprich J, et al. Measles transmission during a large outbreak in California. Epidemics. 2020;30:100375. doi:10.1016/j.epidem.2019.100375

10. Wallinga J, Teunis P. Different Epidemic Curves for Severe Acute Respiratory Syndrome Reveal Similar Impacts of Control Measures. Am J Epidemiol. 2004;160(6):509–516. doi:10.1093/aje/kwh255

11. Patel MK, Antoni S, Nedelec Y, et al. The Changing Global Epidemiology of Measles, 2013–2018. J Infect Dis. 2020;222(7):1117–1128. doi:10.1093/infdis/jiaa044

12. HгyeH MЧ, HгyeH MФ, Лe MX, et al. Epidemiological manifestations of measles in Vietnam and the Russian Federation in 2000–2024. Epidemiol Vaccin. 2026;25(2):29–38. doi:10.31631/2073-30462026-26-2-29-38

13. Robert A, Suffel AM, Kucharski AJ. Long-term waning of vaccine-induced immunity to measles in England: a mathematical modelling study. Lancet Public Health. 2024;9(10):e766–e775. doi:10.1016/S2468-2667(24)00181-6

14. Mehra S, Kludkleeb S, Chaimayo C, et al. Unveiling immunity gaps and determining a suitable age for a third dose of the measles-containing vaccine: a strategic approach to accelerating measles elimination. Lancet Reg Health - Southeast Asia. 2025;32. doi:10.1016/j.lansea.2024.100523

